# A model to predict COVID-19 epidemics with applications to South Korea, Italy, and Spain

**DOI:** 10.1101/2020.04.07.20056945

**Authors:** Z. Liu, P. Magal, Ousmane Seydi, Glenn Webb

## Abstract

In this work, our team develops a differential equations model of COVID-19 epidemics. Our goal is to predict forward in time the future number of cases from early reported case data in regions throughout the world. Our model incorporates the following important elements of COVID-19 epidemics: (1) the number of asymptomatic infectious individuals (with very mild or no symptoms), (2) the number of symptomatic reported infectious individuals (with severe symptoms) and (3) the number of symptomatic unreported infectious individuals (with less severe symptoms). We apply our model to COVID-!9 epidemics in South Korea, Italy and Spain.

## 2 Model

In previous works [1], [2], [3], our team developed differential equations models of COVID-19 epidemics. Our goal was to predict forward in time the future number of cases from early reported case data in regions throughout the world. Our models incorporated the following important elements of COVID-19 epidemics: (1) the number of asymptomatic infectious individuals (with very mild or no symptoms), (2) the number of symptomatic reported infectious individuals (with severe symptoms) and (3) the number of symptomatic unreported infectious individuals (with less severe symptoms). Our models decomposed COVID-19 epidemics into three phases:

Phase I: *the number of cumulative reported cases increases linearly day by day;*

Phase II: *the number of cumulative reported cases increases exponentially day by day;*

Phase III: *the number of daily reported cases decreases day by day*.

The transitions between phases are generally difficult to determine, but can be estimated from reported cases data, as time progresses.

Our model here consists of the following differential equations and initial conditions:

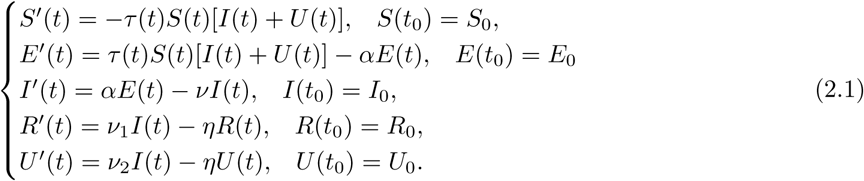

Here *t* ≥ *t*_0_ is time in days, *t*_0_ is the beginning date of the epidemic, *S*(*t*) is the number of individuals susceptible to infection at time *t, E*(*t*) is the number of asymptomatic noninfectious (exposed or latent infected) individuals at time *t, I*(*t*) is the number of asymptomatic but infectious individuals at time *t, R*(*t*) is the number of reported symptomatic infectious individuals at time *t*, and *U*(*t*) is the number of unreported symptomatic infectious individuals at time *t*.

The time-dependent transmission rate parameter is *τ*(*t*). Newly infected noninfectious asymptomatic individuals *E*(*t*) are incubating an average period of 1/*α* days. Asymptomatic infectious individuals *I*(*t*) are infectious for an average period of 1/*ν* days. Reported symptomatic infectious individuals *R*(*t*) are infectious for an average period of 1/*η* days, as are unreported symptomatic infectious individuals *U*(*t*). We assume that reported symptomatic infectious individuals *R*(*t*) are reported and isolated immediately, and cause no further infections. The asymptomatic individuals *I*(*t*) can also be viewed as having a low-level symptomatic state. All infections are acquired from either *I*(*t*) or *U*(*t*) infectious individuals. The fraction *f* of asymptomatic infectious become reported symptomatic infectious, and the fraction 1 − *f* become unreported symptomatic infectious. The rate at which asymptomatic infectious become reported symptomatic is *ν*_1_ = *f ν*, the rate at which asymptomatic infectious become unreported symptomatic is *ν*_2_ = (1 − *f*) *ν*, where *ν*_1_ + *ν*_2_ = *ν*.

The cumulative number of reported cases *CR*(*t*) at time *t* is

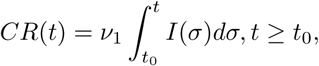

the cumulative number of unreported cases *CU*(*t*) at time *t* is

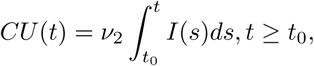

and the daily number of reported cases *DR*(*t*) at time *t* is obtained from the solution of the equation

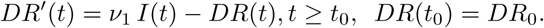

A flow diagram of the model is given in Figure 1.

**Figure 1:**
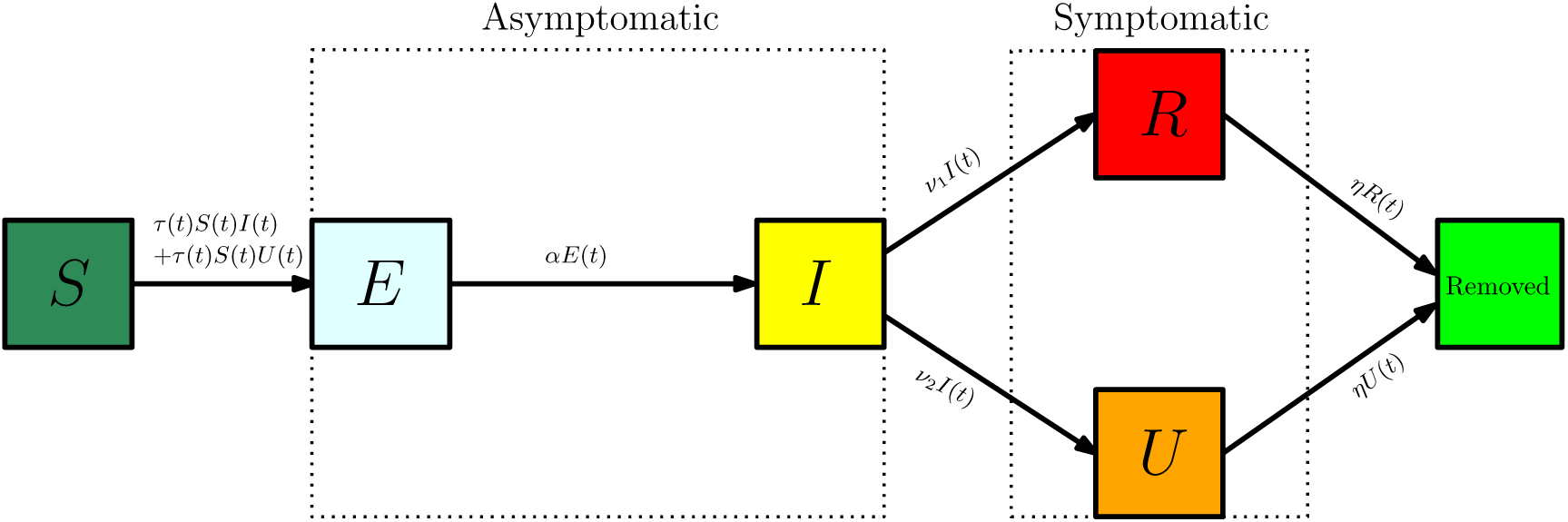
Compartments and flow chart of the model.

**Figure 2:**
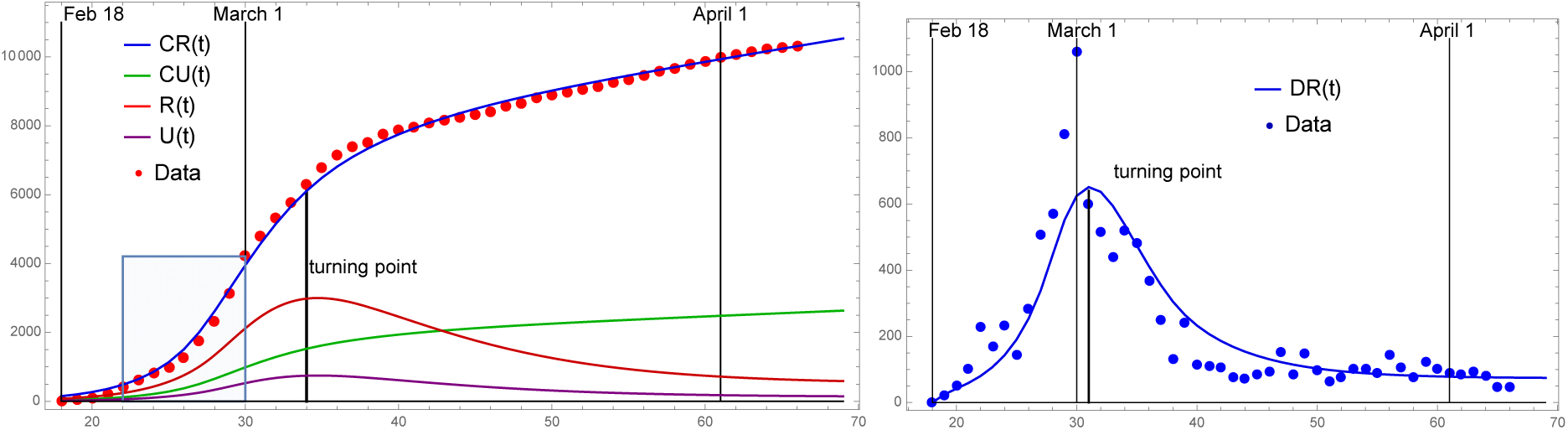
Model simulation for South Korea: Left side - cumulative reported cases: shaded region = Phase II, model turning point = March 5. Right side - daily reported cases: model turning point = March 2.

**Figure 3:**
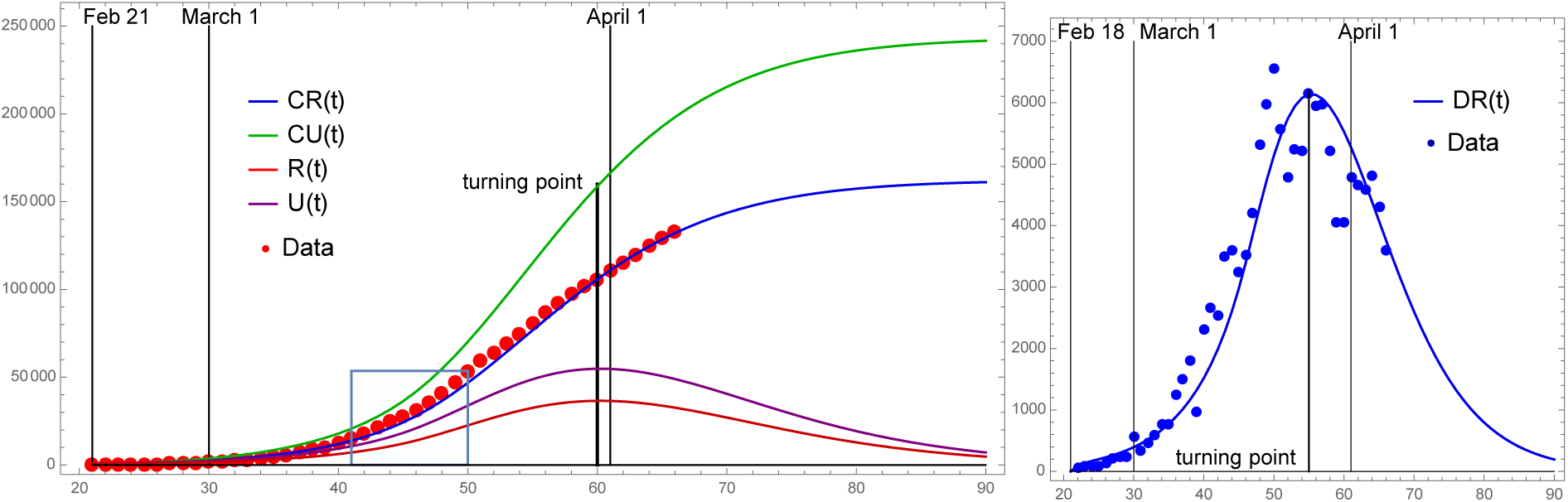
Model simulation for Italy: Left side - cumulative reported cases: shaded region = Phase II, model turning point = March 31. Right side - daily reported cases: model turning point = March 26.

**Figure 4:**
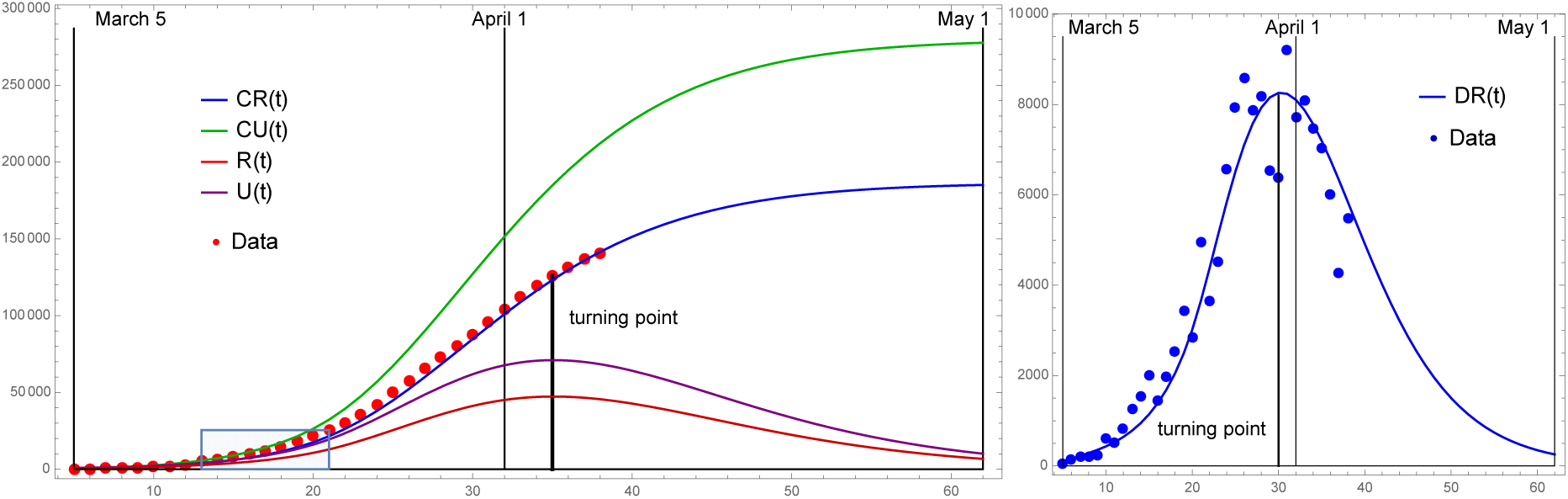
Model simulation for Spain: Left side - cumulative reported cases: shaded region = Phase II, model turning point = April 4. Right side - daily reported cases: model turning point = March 30.

## 3 Parameters

The fraction *f* of total symptomatic infectious cases that are reported is unknown, and varies from region to region. We assume *η* = 1/7, which means that the average period of infectiousness of both unreported symptomatic infectious individuals and reported symptomatic infectious individuals is 7 days. We assume *ν* = 1/6, which means that the average period of infectiousness of asymptomatic infectious individuals is 6 days. We assume *α* = 1, which means that the average period of exposed individuals is 1 day. These values can be modified as further epidemiological information becomes known. At present, they are consistent with accepted values.

A COVID-19 epidemic transitions from Phase I to Phase II at the time *t*_1_ > *t*_0_. Before *t*_1_ the cumulative number of reported cases data increases linearly day by day. After *t*_1_ the cumulative reported cases data increases exponentially day by day. The value of *t*_1_ is estimated from the cumulative reported cases data. We fit an exponentially growing curve *CR*(*t*) to the cumulative reported cases data in an estimated time interval [*t*_1_, *t*_2_], according to the formula:

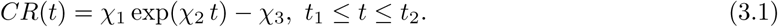

We typically set the value *χ*_3_ = 1, but allow for other values. The initial value *S*_0_ corresponds to the population of the region of the reported case data. The other initial conditions are

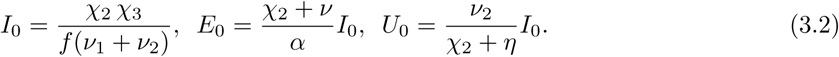

Further, the value of *t*_0_ (when *R*(*t*_0_) = *CR*(*t*_0_) = 0) for the starting time *t*_0_ of the epidemic in the model is given by

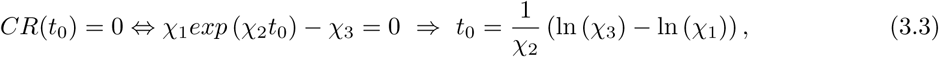

Additionally,

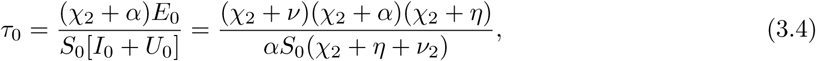

and the basic reproductive number is given by

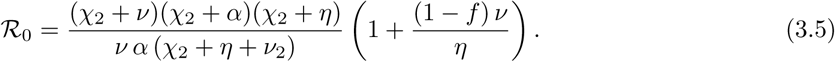

These formulas for *I*_0_, *E*_0_, *U*_0_, *t*_0_, *τ*_0_, and ℛ_0_ were derived in [3]. Their values connect the Phase II reported cases data to the parameterisation and initialisation of our differential equations model.

During Phase II of the epidemic, *τ*(*t*) ≡ *τ*_0_ is constant. When strong government measures such as isolation, quarantine, and public closings are implemented, Phase III begins. The timing of the implementation of these measures, and their impact on disease transmission, is complex. We use an exponentially decreasing time-dependent transmission rate *τ*(*t*) in Phase III to incorporate these effects. The formula for *τ*(*t*), which includes Phase III beginning on day *N*, is

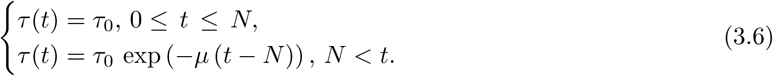

The date *N* and the intensity *µ* of the public measures are chosen so that the cumulative reported cases in the numerical simulation of the epidemic aligns with the cumulative reported case data at an identified date after day *N*. In this way, we are able to project forward the time-path of the epidemic after the government imposed public measures take effect.

## 4 Applications

We apply our model to South Korea, Italy, and Spain ([4, 5, 6]). In Table 1 we provide the parameters for these three countries.

**Table 1:**
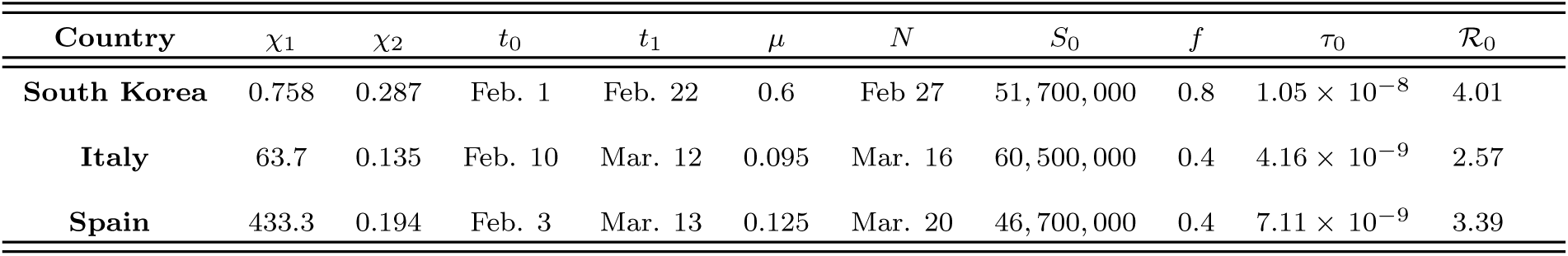
The parameters *χ*_1_, *χ*_2_ are obtained by fitting *χ*_1_ exp(*χ*_2_ *t*) − 1.0 to the cumulative reported cases data between the dates [*t*_1_, *t*_2_] for each country:(1) *t*_1_ = February 22 to *t*_2_ = March 1 for South Korea;(2) *t*_1_ = March 12 to *t*_2_ = March 21 for Italy;(3) *t*_1_ = March 13 to *t*_2_ = March 21 for Spain. The values of *I*_0_ *U*_0_, *τ*_0_, *t*_0_, *τ*_0_, and ℛ_0_ are obtained by using(3.2), (3.3), (3.4), (3.5). The parameters *ν* = 1/6, *η* = 1/7, *α* = 1/1, *χ*_3_ = 1.0, and *R*_0_ = 1.0 for all three countries.

| Country | $\chi_1$ | $\chi_2$ | $t_0$ | $t_1$ | $\mu$ | $N$ | $S_0$ | $f$ | $\tau_0$ | $\mathcal{R}_0$ |
| --- | --- | --- | --- | --- | --- | --- | --- | --- | --- | --- |
| South Korea | 0.758 | 0.287 | Feb. 1 | Feb. 22 | 0.6 | Feb 27 | 51,700,000 | 0.8 | $1.05 \times 10^{-8}$ | 4.01 |
| Italy | 63.7 | 0.135 | Feb. 10 | Mar. 12 | 0.095 | Mar. 16 | 60,500,000 | 0.4 | $4.16 \times 10^{-9}$ | 2.57 |
| Spain | 433.3 | 0.194 | Feb. 3 | Mar. 13 | 0.125 | Mar. 20 | 46,700,000 | 0.4 | $7.11 \times 10^{-9}$ | 3.39 |

### 4.1 COVID-19 epidemic in South Korea

The epidemic in South Korea can be divided into four stages:

1. Before February 22: Phase I.
2. February 22 to March 1: Phase II.
3. March 2 to March 8: Phase III. The South Korean government implemented extensive testing, isolation, and contact tracing of confirmed cases, and quarantine policies after February 20, which took effect in daily reported cases after March 2.
4. After March 8: The daily reported cases were approximately the same each day and the cumulative reported cases increased linearly. This stage corresponds to a new Phase I, with a low level background generation of reported cases each day. To account for this new Phase I, the model (2.1) is modified by replacing *τ*(*t*) in (2.1) with a new transmission function *τ*(*t, S*(*t*), *I*(*t*), *U*(*t*)) that depends on *t, S*(*t*), *I*(*t*), *U*(*t*) as follows:

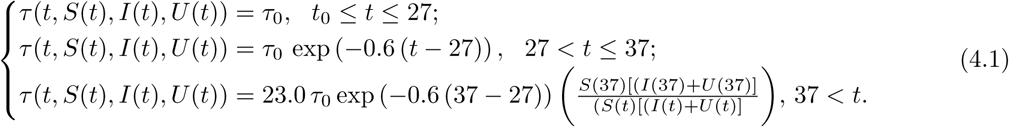

The value 23.0 is chosen to match the slope of the linear increasing cumulative reported cases data after day 37. The equations and initial values are the same, except for this new *τ* function. The formulas in (4.1) connect the new Phase I to the transmission rate in the model equations, and to the model outputs of *E*(*t*), *I*(*t*), *U*(*t*), *R*(*t*), *CU*(*t*), *CR*(*t*), *DR*(*t*). The form of (4.1) can be applied to other examples which transition from Phase III to a new Phase I, corresponding to a linearly increasing growth rate of cumulative reported cases. This new Phase I can further transition to another Phase I with slower linearly increasing growth rate,

### 4.2 COVID-19 epidemic in Italy

The epidemic in Italy can be divided into three stages:

1. Before March 12: Phase I.
2. March 12 to March 21: Phase II.
3. Beginning March 1, the Italian government implemented extensive public regional lockdown measures, which were extended to all of Italy on March 10. These measures took effect in reducing reported daily cases approximately two weeks later. We take Phase III to be March 24 onward.

### 4.3 COVID-19 epidemic in Spain

The epidemic in Spain can be divided into three stages:

1. Before March 13: Phase I.
2. March 13 to March 21: Phase II.
3. On March 13, the Spanish government implemented partial shutdown measures, and on March 14 imposed a general state of alarm on all of Spain. These measures took effect in reducing reported daily cases approximately two weeks later. We take Phase III to be March 28 onward.

## 5 Conclusions

We have applied a method developed in [1], [2], [3] to predict the evolution of a COVID-19 epidemic in a geographical region, based on reported case data in that region. Our model focuses on unreported cases, asymptomatic infectious cases, and the division of the epidemic evolution through a succession of phases. Our method can be predictive, when the epidemic is growing exponentially in Phase II. In [1] we demonstrated a method to identify the Phase II exponentially increasing rate of cumulative reported cases. When public measures are begun in Phase II, to ameliorate the epidemic, we model these measures with a time-dependent exponentially decreasing transmission rate. These measures result in a subsequent reduction in daily reported cases, which we call Phase III. We determine the transition from Phase II to Phase III, which may require more than a week, in the model simulations.

For South Korea, the epidemic has attenuated, because of the major measures that were implemented to restrict public distancing. These measures involved surveillance, extensive testing, isolation and contact tracing of reported suspected andcases. The cumulative number of reported cases in South Korea, however, has not flattened, but rather is growing linearly at a low rate. For Italy and Spain, the epidemics have evidently passed the turning point, according to the daily reported cases data. The cumulative reported cases may not flatten, but as in South Korea, will continue to grow linearly at a low rate.

Our model incorporates government and social distancing measures, through the time-dependent transmission rate *τ*. It is evident that these measures should start as early as possible, and should be as strong as possible. If the epidemic subsides substantially due to these measures, the example of South Korea shows that a background level of daily cases may persist for an extended time. If major distancing measures are reduced too early or too extensively, the epidemic may return to new Phase II, with exponentially increasing cumulative cases. A possible control of COVID-19 epidemics is evidenced by the example of South Korea. The future of COVID-19 epidemics and their human toll is at present uncertain, and it is hopeful that mathematical models can be of use.

## Data Availability

All data used is available through the web site wikipedia as listed in our references.

https://en.wikipedia.org/wiki/2020_coronavirus_outbreak_in_South_Korea

https://en.wikipedia.org/wiki/2020_coronavirus_outbreak_in_Italy

https://en.wikipedia.org/wiki/2020_coronavirus_outbreak_in_Spain

## References

[1] Z. Liu, P. Magal, O. Seydi, and G. Webb, Understanding unreported cases in the 2019-nCov epidemic outbreak in Wuhan, China, and the importance of major public health interventions, MPDI Biology, 2020, 9(3), 50.

[2] Z. Liu, P. Magal, O. Seydi, and G. Webb, Predicting the cumulative number of cases for the COVID-19 epidemic in China from early data, medRxiv, 2020.

[3] Z. Liu, P. Magal, O. Seydi, and G. Webb, A COVID-19 epidemic model with latency period, to appear.

[4] https://en.wikipedia.org/wiki/2020_coronavirus_outbreak_in_South_Korea

[5] https://en.wikipedia.org/wiki/2020_coronavirus_outbreak_in_Italy

[6] https://en.wikipedia.org/wiki/2020_coronavirus_outbreak_in_Spain

